# SARS-Cov-2 cysteine-like protease (Mpro) is immunogenic and can be detected in serum and saliva of COVID-19-seropositive individuals

**DOI:** 10.1101/2020.07.16.20155853

**Authors:** Pedro Martínez-Fleta, Arantzazu Alfranca, Isidoro González-Álvaro, Jose M Casasnovas, Daniel Fernández-Soto, Gloria Esteso, Yaiza Cáceres-Martell, Sofía Gardeta, Salomé Prat, Tamara Mateu-Albero, Ligia Gabrie, Eduardo López-Granados, Francisco Sánchez-Madrid, Hugh T. Reyburn, José M. Rodríguez Frade, Mar Valés-Gómez

## Abstract

Currently, there is a need for reliable tests that allow identification of individuals that have been infected with SARS-CoV-2 even if the infection was asymptomatic. To date, the vast majority of the serological tests for SARS-CoV-2 specific antibodies are based on serum detection of antibodies to either the viral spike glycoprotein (the major target for neutralising antibodies) or the viral nucleocapsid protein that are known to be highly immunogenic in other coronaviruses. Conceivably, exposure of antigens released from infected cells could stimulate antibody responses that might correlate with tissue damage and, hence, they may have some value as a prognostic indicator. We addressed whether other non-structural viral proteins, not incorporated into the infectious viral particle, specifically the viral cysteine-like protease, might also be potent immunogens. Using ELISA tests, coating several SARS-CoV-2 proteins produced in vitro, we describe that COVID-19 patients make high titre IgG, IgM and IgA antibody responses to the Cys-like protease from SARS-CoV-2, also known as 3CLpro or Mpro, and it can be used to identify individuals with positive serology against the coronavirus. Higher antibody titres in these assays associated with more severe disease and no cross-reactive antibodies against prior betacoronavirus were found. Remarkably, IgG antibodies specific for Mpro and other SARS-CoV-2 antigens can also be detected in saliva. In conclusion, Mpro is a potent antigen in infected patients that can be used in serological tests and its detection in saliva could be the basis for a rapid, non-invasive test for COVID-19 seropositivity.

## INTRODUCTION

The identification of the link between a novel beta-coronavirus strain, named Severe Acute Respiratory Syndrome-CoronaVirus-2 (SARS-CoV-2), and a fatal respiratory illness, COVID-19, formally recognised as a pandemic by the WHO on March 11 (1, 2) has led to a rush by health systems all over the world to develop and implement testing for viral infection. The rapid cloning and sequencing of the viral genome permitted the development of PCR-based assays for the detection of viral nucleic acids that have become a key strategy for both clinical diagnosis and epidemiological monitoring studies. However, besides identifying individuals with active infection, it is also necessary to know which patients, either symptomatic or asymptomatic, have developed an antibody response to the virus. Several reasons make SARS-CoV-2 serology tests crucial. First, PCR testing is not 100% efficient, (3–5). Second, testing for viral RNA cannot detect evidence of past infection, which will be crucial for epidemiological efforts to assess how many people have been infected in any given area. In addition, this will allow definition of the infection fatality rate and help with management of the epidemic. Third, assays to measure antibody responses and determine seroconversion, while not appropriate to detect acute infections, are however, valuable sources of information on the quality of the response exerted by different individuals developing different clinical manifestations. Moreover, if different isotypes and viral antigens are included in assays testing different time points after the onset of the disease, information of clinical importance will be produced. Finally, quantitative and qualitative assays of antibody responses can aid in the identification of factors that correlate with effective immunity to SARS-CoV-2, the duration of these immune responses and may also aid in the selection of donors from whom preparations of convalescent serum/plasma can be generated for therapeutic use.

Multiple antibody tests to detect exposure to SARS-CoV-2, are becoming available. The majority of these assays have been optimised to detect immunoglobulin G (IgG) and, in some cases, IgM antibodies using different viral antigens, being the Spike (S) protein and the nucleoprotein of SARS-CoV-2 the more widely used (6, 7). These proteins are key elements of the viral particle and are expected, by analogy with other coronaviruses, to be highly immunogenic. However, the immunogenicity of other viral proteins, 28 are encoded in the viral genome, has been little explored. Here we have studied the antibody response to the main viral protease (Mpro, or 3CLPro) elicited after viral infection. Although this protein is not exposed in the viral particle, Mpro carries out a critical role in viral replication. Like other beta-coronaviruses, SARS-CoV-2 is a positive-sense RNA virus that expresses all of its proteins as a single polypeptide chain and Mpro cleaves the 1ab polyprotein to yield the rest of the mature proteins of the virus. Since this activity is essential for the viral life cycle, Mpro structure and function has been studied intensively (8); in particular, Mpro has been suggested as a target for specific inhibitors that might act as potent anti-viral agents (9). However, to our knowledge, no study on the antigenicity of this protease has been reported.

To increase the possibilities of diagnosing COVID-19 patients, here we report the use of an ELISA test involving the assay of sero-reactivity to three different SARS-CoV-2 antigens, including the protease Mpro. These data demonstrate that individuals who have been infected with SARS-CoV-2 make high titre antibody responses to Mpro and that assays for seroreactivity to this protein sensitively and specifically discriminate between infected and non-infected individuals. Further, while most available tests assess for SARS-CoV-2-specific IgM and IgG antibodies, here, we also explored the presence of IgA antibodies in the sera tested. While, in general, assays for IgM antibodies resulted in a high background that limited the sensitivity of the ELISA, testing for IgA seropositivity provided very clean data, with low background and high signal, therefore providing a very good tool to complement IgG assays.

Interestingly, considerable significant amounts of IgA antibodies specific for MPro, as well as the Receptor Binding Domain (RBD) and NP, were also frequently found in serum of COVID-19 infected individuals and the amounts of IgA and IgM antibodies could be related with disease severity.

Surprisingly, IgG antibodies specific for SARS-CoV-2 antigens were also readily detectable in the saliva of these patients and, in this case, the titre of protease-specific antibodies was higher than for the other two proteins tested. Since the nasal and buccal mucosa are key sites of viral infection and replication, the presence of antibodies in saliva may be an important feature of the virus-specific immune response, but this observation may also allow the development of a rapid, completely non-invasive assay for COVID-19 seropositivity.

## RESULTS

### Mpro-specific antibodies can be detected in serum of COVID-19 patients by ELISA

Since this study evaluated, for the first time, whether coronavirus-infected individuals could generate an antibody response against the Cys-like protease, MPro, other SARS-CoV-2 proteins, commonly used in serology tests, were produced, for comparison. Mpro and NP were expressed in *E coli*, and two different constructs of the Receptor Binding Domain (RBD) of the spike protein were used: one was expressed by transfection in mammalian cells (mRBD) and a second, produced by baculovirus infection of insect cells (iRBD-His). All the proteins, except mRBD, had a histidine-tag and they were purified on Ni^2+^-NTA columns followed by size exclusion chromatography (Figure 1).

**Figure 1.**
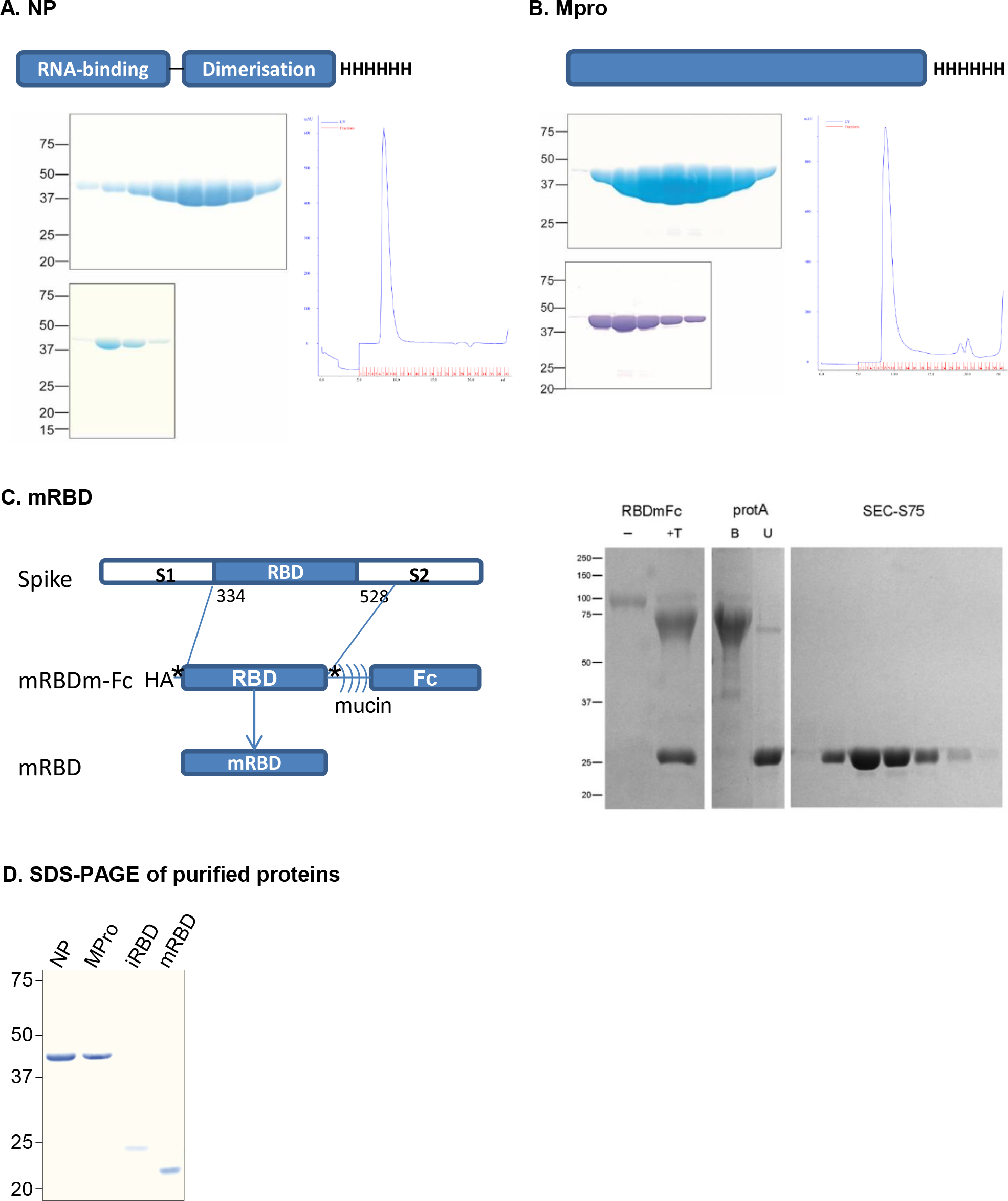
SARS-CoV-2 protein purification. Nucleocapsid (NP) (**A**) and Cys-like protease (3CLpro, Mpro) (**B**) proteins were expressed in E. coli and extracted from the soluble fraction of the bacterial pellet. The proteins were firstly purified by selection through their His-tags in HiTrap Ni2+ chelating columns. The fractions eluted from these columns were run in SDS-PAGE (top gels). After that, proteins were further purified by gel filtration using a Superdex 75 column and fractions eluted from this step were run in SDS-PAGE (bottom gels). The FPLC profile is shown on the right panels. (**C**) mRBD The 334-528 fragment of the Spike protein was produced in mammalian cells fused to an HA-tag, at the N-terminus and to the TIM-1 mucin domain followed by the Fc portion of human IgG, at the C-terminus. Two thrombin-recognition sites (asterisks) were introduced. The fusion protein was treated with thrombin (+T in the SDS-PAGE shown at the right) to release the mRBD fragment. It was further purified using a protein A column and size exclusion chromatography (Superdex 75). SDS-PAGE under reducing conditions are shown for the samples at the purification steps. Proteins bound (B) and unbound (U) to the protein A column are shown. (**D**) SDS-PAGE. After expression in the different systems, proteins were purified and fractions from gel filtration chromatography were run in SDS-PAGE under non reducing conditions.

Before testing a large number of sera from COVID-19 patients and healthy donors, experiments were designed to optimize coating and dilution conditions. These data already revealed that COVID-19 patient sera contained high titres of Mpro-specific antibodies. Antibody reactivity to the viral protease reached saturation at relatively low concentrations and discriminated efficiently between individuals who had been infected with SARS-CoV-2 and those that had not been exposed to the virus (Figure 2A). Serum dilutions from 1/50 to 1/1600 covered a broad range of reactivity to Mpro from almost no recognition to saturation (reached at 1/100 dilution). It was also possible to detect low titres of antibodies of the IgM and IgA isotypes in these patients (Figure 2B), suggesting that, in subsequent experiments, a large screening of patient samples should be performed including the three Ig subclasses. Coating titration experiments further confirmed the specificity of the assay (Figure 2C). The IgG reactivity against the protease MPro in COVID-19 patients was comparable, or in certain cases stronger, to the reactivity against RBD, however, no differences were noticed between the RBD recombinant proteins expressed in either mammalian cells or baculovirus (Supplementary Figure 1). These initial experiments suggested that the humoral response against the three viral proteins can be heterogeneous between different patients.

**Figure 2.**
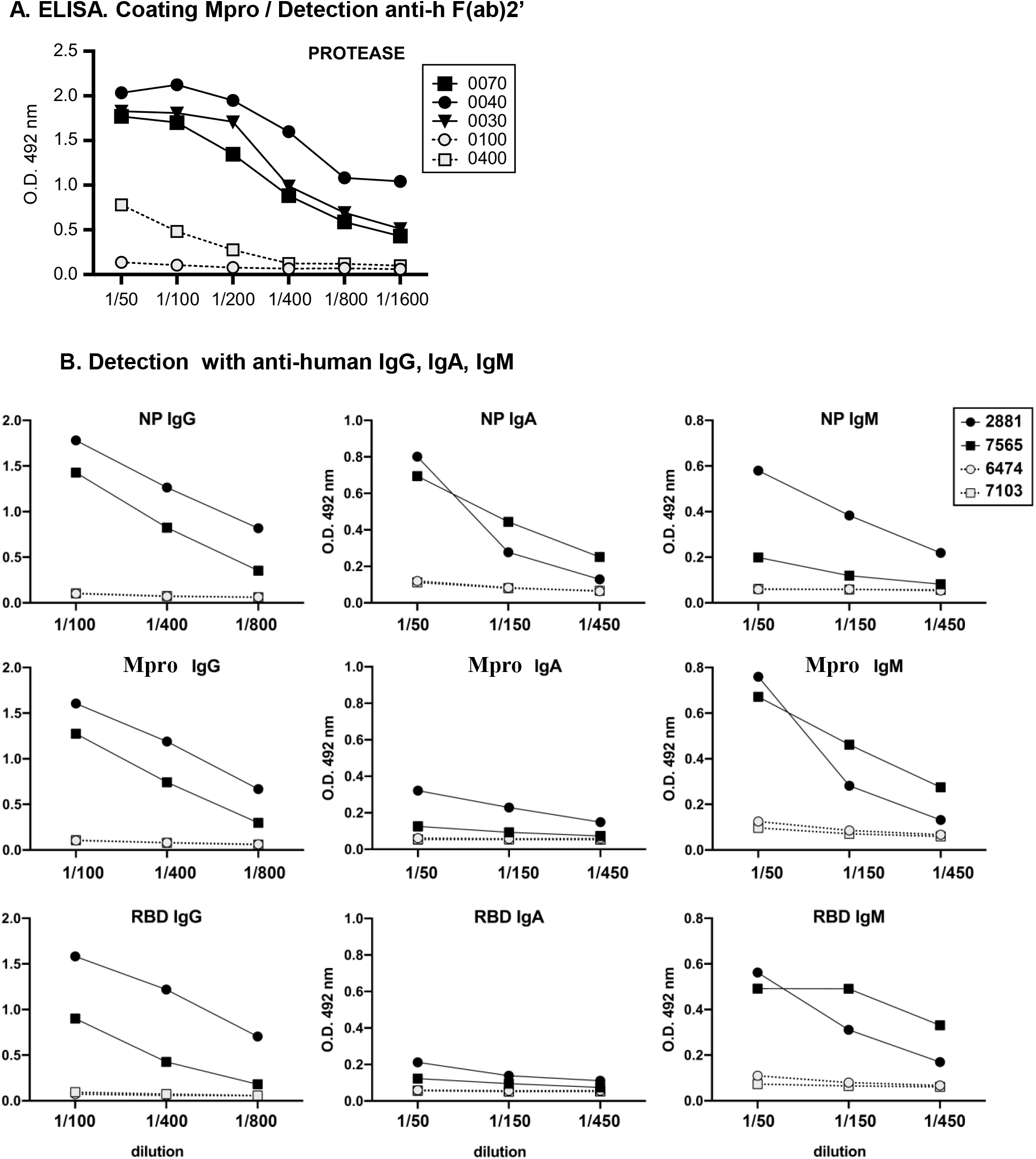
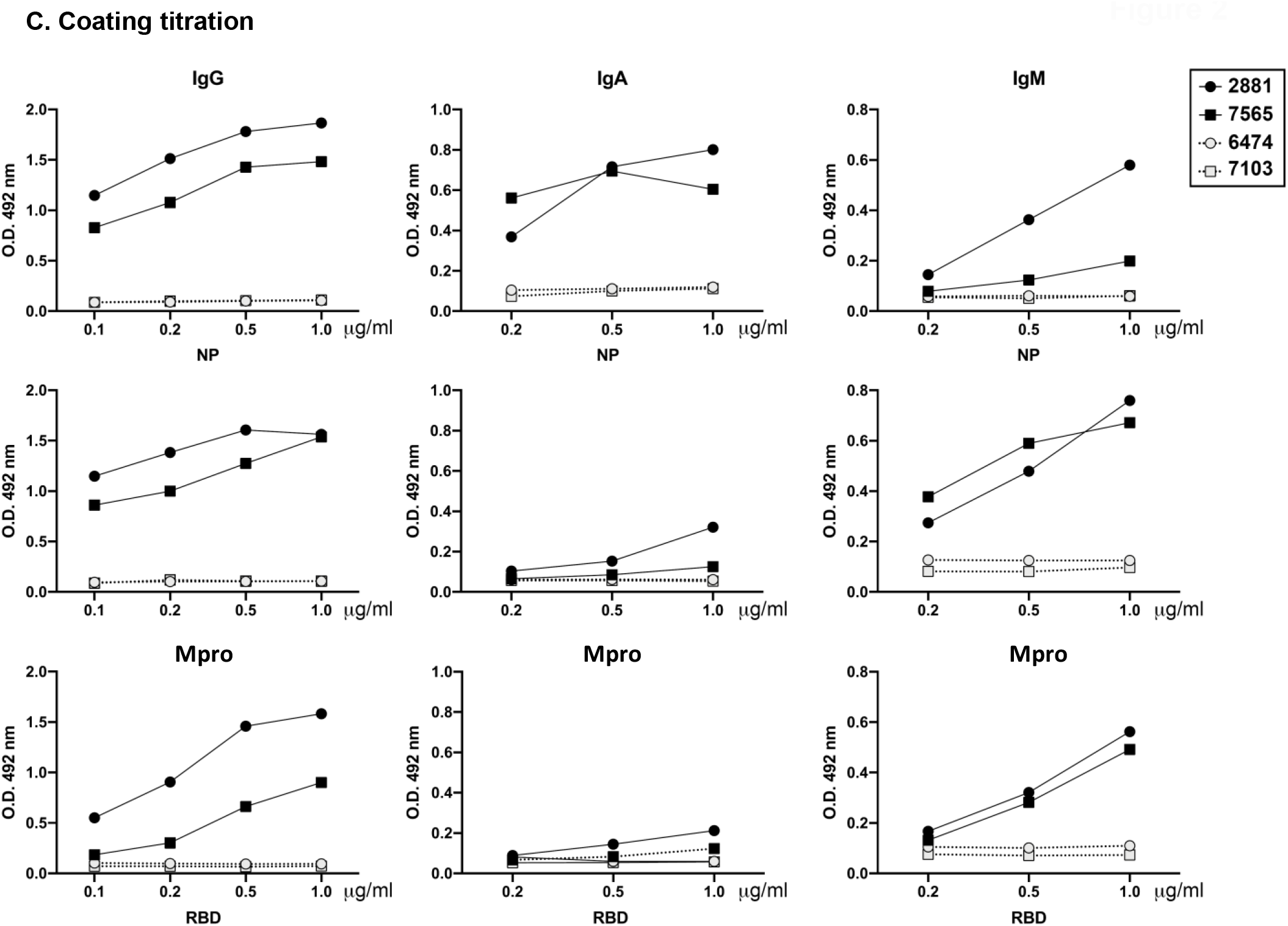
Detection of SARS-CoV-2 Mpro-specific antibodies by ELISA. (A) Sera titration on Mpro. Plates were coated with SARS-CoV-2 Mpro and sera dilutions (1/50 to 1/1600) were tested. Detection was performed using anti-human F(ab)2’ antibody. **(B) Isotype recognition**. Plates coated with SARS-CoV-2 Mpro, nucleoprotein (NP) and RBD were detected with antibodies directed against human Ig of the three different subclasses: IgG, IgA, IgM. Black symbols correspond to COVID-19 patients and grey symbols to donors pre-COVID-19. (**C**) **Coating titration**. Plates were coated with increasing amounts of SARS-CoV-2 Mpro, nucleoprotein (NP) and RBD and sera diluted 1/100 for IgG detection and 1/50 for IgA and IgM were tested. Black symbols correspond to COVID-19 patients and grey symbols to donors pre-COVID-19.

To further validate the assay, additional controls were performed such as monitoring the background in plates with no viral antigen coating and testing sera collected before the COVID-19 pandemic (Supplementary Figure 2).

### Detection of SARS-CoV-2 Mpro-specific antibodies identifies COVID-19 seropositive individuals with high specificity and sensitivity

A cohort of 36 COVID-19 patients (PCR+) and 33 healthy donors was recruited at La Princesa University Hospital, Madrid (Table 1) and ELISA assays were performed to detect Mpro-, as well as RBD- and NP-, specific antibodies of the IgG, IgA and IgM subclasses in sera (Figure 3).

**Table 1.** Patient demographic and clinical data.

|  |  |  | **N=36** | **%** |
| --- | --- | --- | --- | --- |
| Gender | Male |  | 21 | 58 |
|  | Female |  | 15 | 42 |
| Age | < 35 |  | 7 | 19 |
|  | 35-60 |  | 18 | 50 |
|  | > 60 |  | 11 | 31 |
| Time from symptoms onset to sample collection | < 15 days |  | 2 | 6 |
|  | 15-30 days |  | 13 | 36 |
|  | 31-45 days |  | 14 | 39 |
|  | > 45 |  | 7 | 19 |
| Hospitalization | Yes | Ward | 4 | 11 |
|  |  | ICU | 6 | 17 |
|  | No |  | 26 | 72 |
| Fever |  |  | 31 | 86 |
| Shivers |  |  | 23 | 64 |
| Headache |  |  | 22 | 61 |
| Confusion |  |  | 6 | 17 |
| Conjunctival congestion |  |  | 5 | 14 |
| Nasal congestion |  |  | 18 | 50 |
| Rhinorrhea |  |  | 16 | 44 |
| Anosmia |  |  | 16 | 44 |
| Ageusia |  |  | 18 | 50 |
| Odynophagia |  |  | 14 | 39 |
| Dry cough |  |  | 19 | 53 |
| Productive cough |  |  | 9 | 25 |
| Dyspnea |  |  | 21 | 58 |
| Chest pain |  |  | 12 | 33 |
| Tonsillitis |  |  | 3 | 8 |
| Adenopathies |  |  | 4 | 11 |
| Nausea/vomiting |  |  | 10 | 28 |
| Diarrhea |  |  | 16 | 44 |
| Skin rash |  |  | 2 | 6 |
| Acrocyanosis |  |  | 1 | 3 |
| Myalgia/arthralgia |  |  | 24 | 67 |
| Asthenia |  |  | 27 | 75 |
| Weight loss |  |  | 20 | 56 |
| Thrombotic events |  |  | 2 | 6 |
| Comorbidities (HTN, DM, COPD, obesity, cancer) |  |  | 17 | 47 |

**Figure 3:**
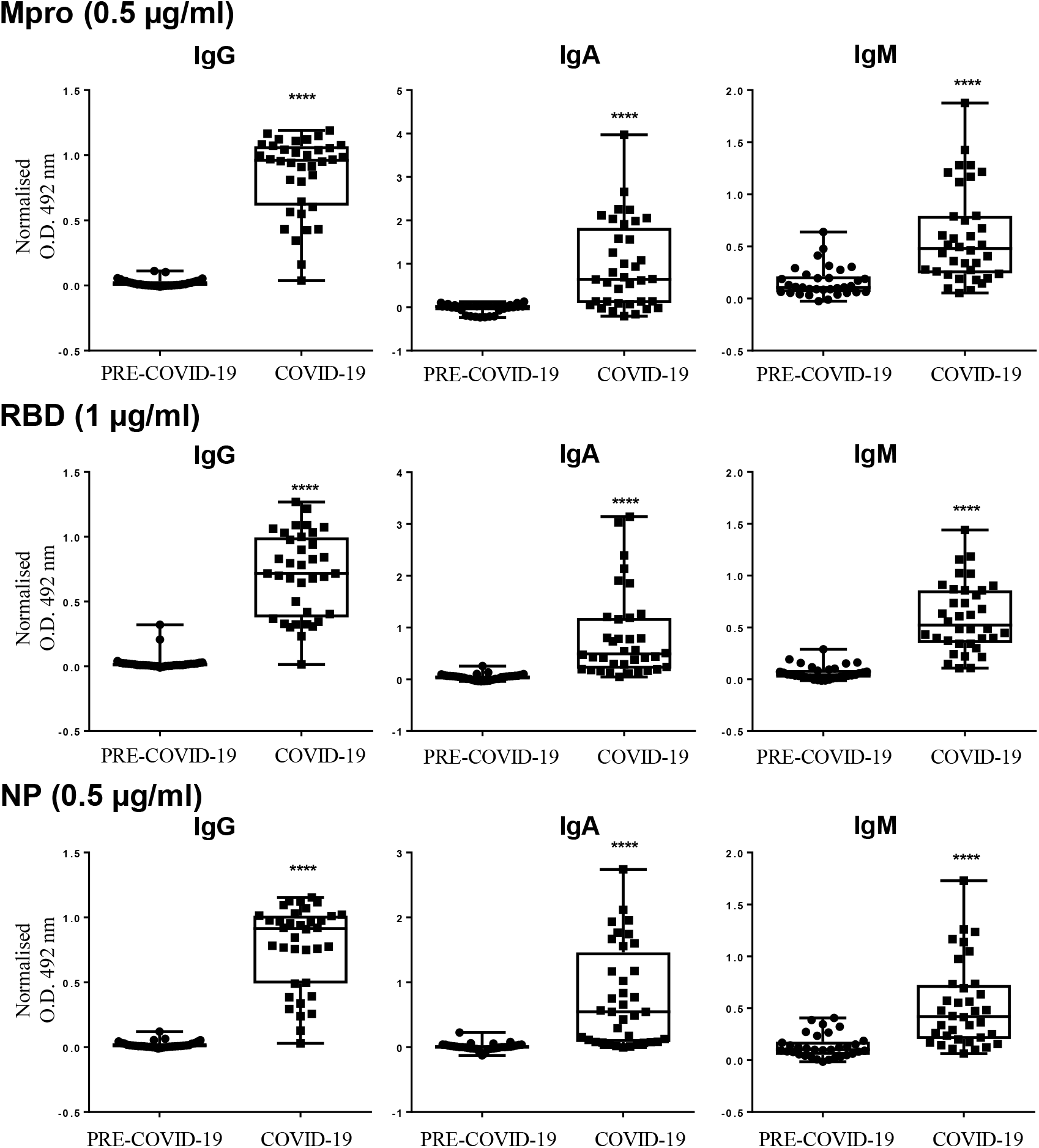
Comparison of sera from 33 pre-COVID-19 vs 36 COVID-19 patients. Plates coated with either 0.5 or 1 µg/ml (as indicated) SARS-CoV-2 Mpro, NP or RBD were used to perform ELISA tests on 36 COVID-19 positive and 33 negative control sera (obtained before the pandemic outbreak, PRE-COVID-19). Detection was done using antibodies directed against human immunoglobulin of the three different subclasses: IgG, IgA, IgM. Sera dilutions from 1/50-1/3200 were carried out. Data were normalised for each antigen using the signal obtained against a pool of positive sera. Box and whisker plots of all the sera tested at the 1/200 dilution for IgG and 1/50 for IgA and IgM. Statistical significance was analysed in Mann-Whitney tests. **** means p<0.0001.

Titration of the serum samples was carried out over a dilution range of 1/50 to1/3200, and these experiments showed that assay for seropositivity to all three antigens discriminated between COVID-19 positive and negative donors, as shown in dot plots comparing different dilutions (Supplementary Figure 3). Figure 3 summarises the absorbance data from all the sera samples. To estimate the cut-off value, the sensitivity, and the specificity parameters for each antigen/Ig isotype pair, receiver operating characteristic (ROC) analyses were performed (Table 2, Figure 4). The best area under the curve (AUC) values were obtained with the measurement of IgG antibodies specific for Mpro and NP (AUCs= 0.9945 and 0.9927, respectively). The sensitivity and specificity was above 90% for detection of IgG antibodies of the three proteins tested, with values of sensitivity and specificity for Mpro of 97% and 100% respectively. AUC values above 0.85 were obtained for the other isotypes (IgA, IgM). Measurement of anti-IgA antibodies appeared to discriminate less accurately between pre-COVID-19 sera and COVID-19 sera, however, this is not due to a lack in sensitivity for this isotype. Instead, because background levels with IgA were very low and the signal clearly positive in some patients, the lack of detection suggests that certain COVID-19-positive patients have circulating IgA while other COVID-19-positive patients lack IgA in peripheral blood. Whether the presence of IgA in periphery has any relationship with clinical aspects needs to be explored further in larger cohorts of patients.

**Table 2.** AUC, cut-off, sensitivity and specificity.

| **Antigen** | **Isotype** | **AUC** | **Cut-off** | **Sensitivity** | **Specificity** |
| --- | --- | --- | --- | --- | --- |
| **RBD** | IgG | 0.961 | 0.232 | 94% | 97% |
|  | IgA | 0.974 | 0.112 | 97% | 94% |
|  | IgM | 0.981 | 0.203 | 91% | 97% |
| **Mpro** | IgG | 0.994 | 0.161 | 97% | 100% |
|  | IgA | 0.833 | 0.130 | 73% | 100% |
|  | IgM | 0.859 | 0.237 | 79% | 79% |
| **NP** | IgG | 0.993 | 0.127 | 97% | 100% |
|  | IgA | 0.949 | 0.066 | 88% | 94% |
|  | IgM | 0.885 | 0.341 | 76% | 85% |

AUC, area under the curve; RBD, Receptor Binding Domain; Mpro, cysteine-like protease; NP, nucleoprotein

**Figure 4:**
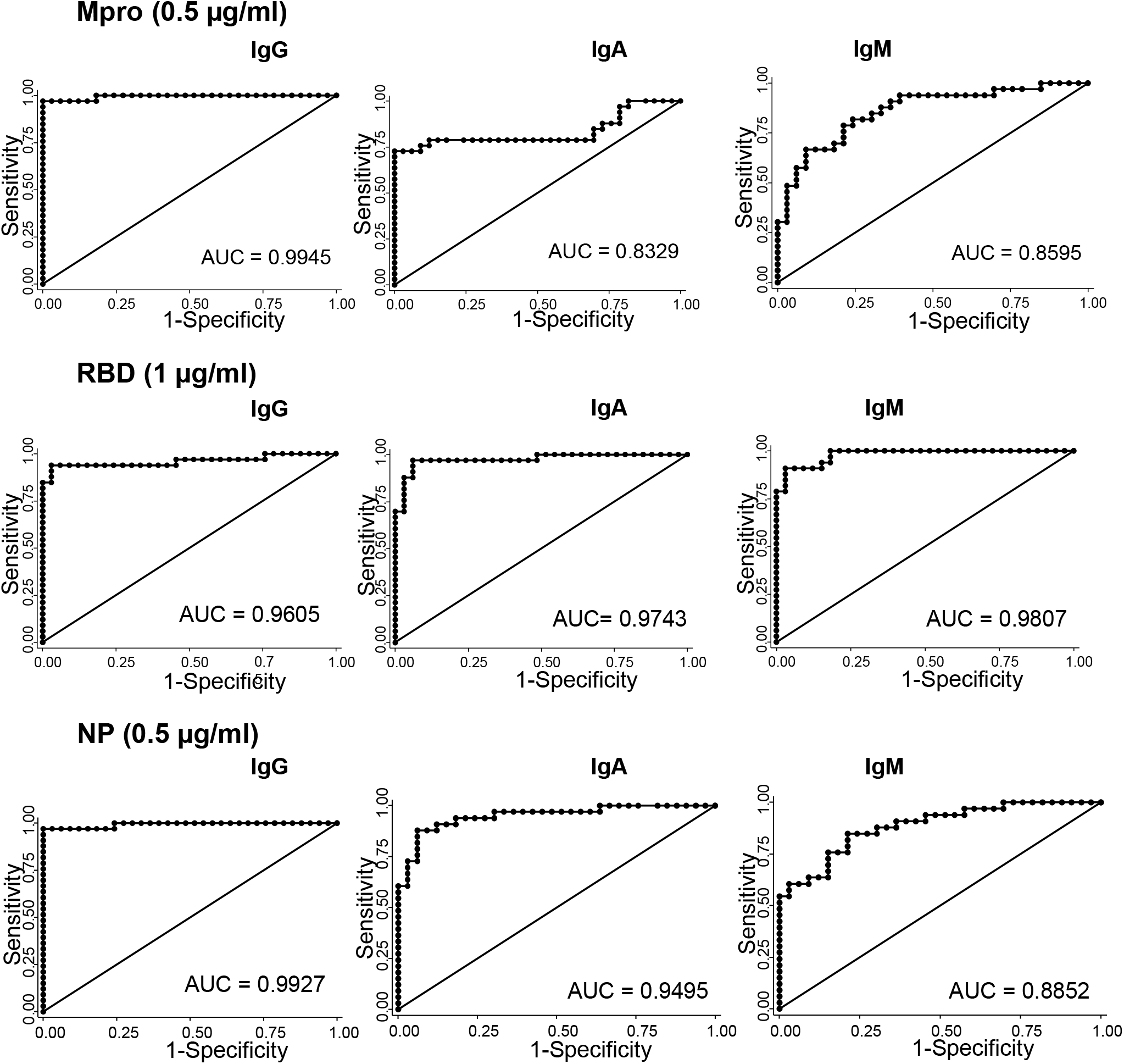
Assessment, through Receiver Operating Characteristic (ROC) analysis, of different isotype responses against three SARS-CoV-2 proteins as COVID-19 classifiers. Graphic representation of the relationship between sensitivity and specificity. The area under the curve (AUC) calculated for each antigen and immunoglobulin pair (see Statistical section of Material and Methods) is indicated. For details on specificity and sensitivity data, see Supplementary Table 1.

Comparison between proteins showed some heterogeneity in the capacity of different donors to produce antibodies, especially for IgM and IgA subclasses. Non-linear polynomial regression showed a better correlation between the detection of antibodies against NP and Mpro compared to NP and RBD or MPro and RBD (Figure 5A). Only one COVID-19 donor failed to make a full antibody response.

**Figure 5.**
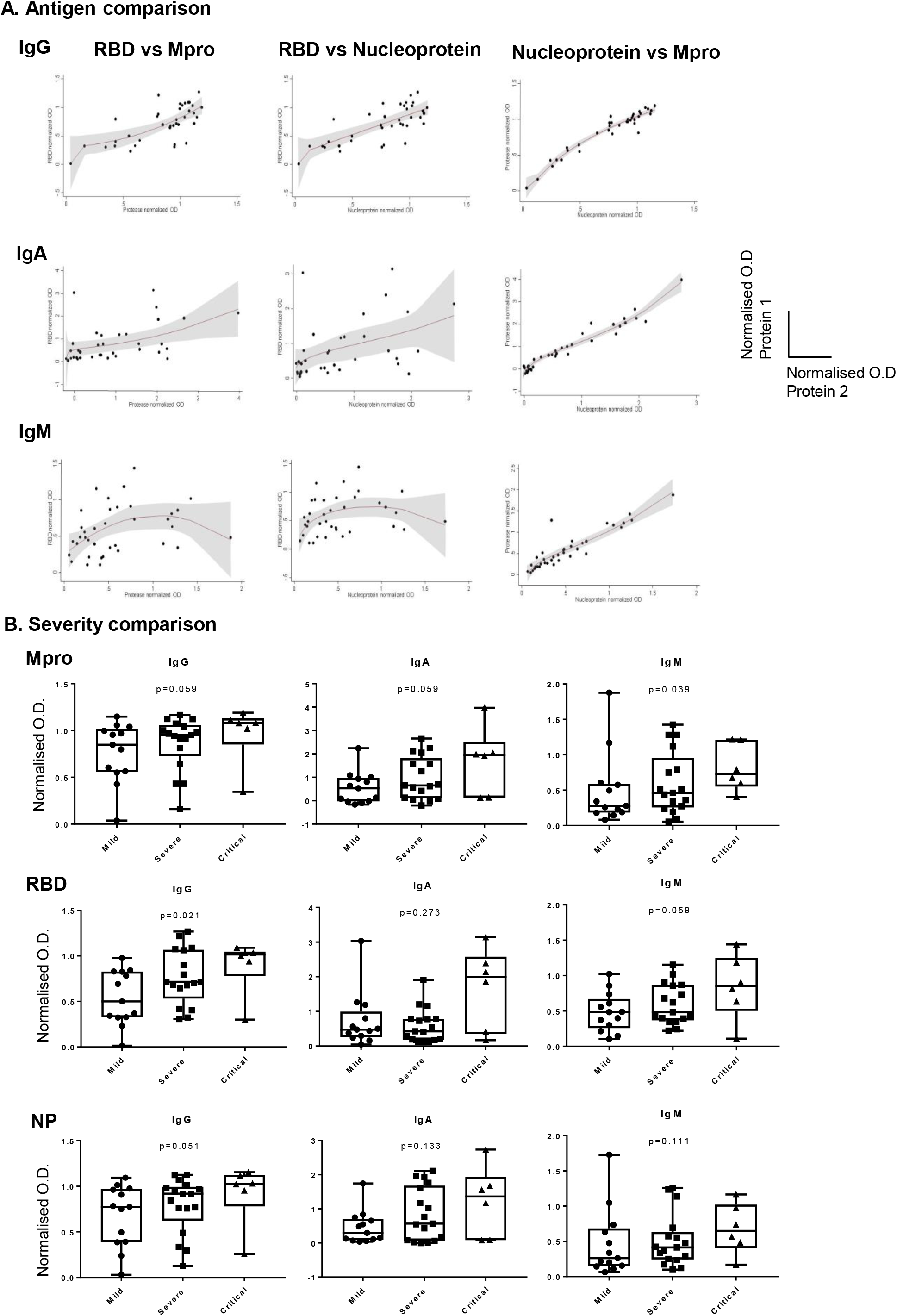
A. Correlations of humoral response against different SARS-CoV-2 antigens by isotype. Data from Figure 2 are shown as dot-plots and their fitted fractional polynomial prediction with 95% confidence interval (transparent grey shadow) estimated using the two-way command of Stata with the fpfitci option. **B. Comparison of sera from mild, severe and critical patients**. Patients were classified into three groups (mild n=13, severe n=17 and critical n=6) according to COVID-19 symptoms severity (see reference 13). Data normalised for each antigen using the signal obtained against a pool of positive sera obtained in Figure 2, are depicted in box and whisker plots at the 1/200 dilution for IgG and 1/50 for IgA and IgM. Statistical significance was analysed by Cuzick’s test.

Further analyses were performed to explore the correlations between the titres of the different antibodies in serum and clinical parameters. Interestingly, a trend for higher titre antibody responses was found in patients with more severe disease (Figure 5B), being more pronounced for IgM against Mpro and IgG against RBD. However, several other variables also contributed to the heterogeneity in antibody response, mainly age and time since the onset of symptoms (Table 3). After adjustment for these possibly confounding factors, IgA anti-RBD was observed to be significantly higher in critical patients compared to patients with mild disease. In addition, critical patients showed a trend to higher IgM and IgA anti-Mpro titres than patients with mild COVID-19. Furthermore, intense IgM and IgA responses against the three proteins were significantly associated with higher serum IL-6 levels (data not shown).

**Table 3.** Variables that explain heterogeneity in antibody response against three proteins of SARS-CoV-2.

|  | Mpro |  |  |  |  |  | RBD |  |  |  |  |  | NP |  |  |  |  |  |
| --- | --- | --- | --- | --- | --- | --- | --- | --- | --- | --- | --- | --- | --- | --- | --- | --- | --- | --- |
|  | IgG |  | IgA |  | IgM |  | IgG |  | IgA |  | IgM |  | IgG |  | IgA |  | IgM |  |
| | $\beta$ Coeff. | p | $\beta$ Coeff. | p | $\beta$ Coeff. | p | $\beta$ Coeff. | p | $\beta$ Coeff. | p | $\beta$ Coeff. | p | $\beta$ Coeff. | p | $\beta$ Coeff. | p | $\beta$ Coeff. | p |
| Age (year) | 0.010 | 0.013 | 0.021 | 0.085 | NRM | - | 0.007 | 0.083 | NRM | - | 0.008 | 0.037 | 0.009 | 0.039 | 0.016 | 0.092 | NRM | - |
| Time since symptoms onset (days) | NRM | - | -0.033 | 0.010 | -0.014 | 0.005 | NRM | - | -0.016 | 0.058 | -0.009 | 0.006 | NRM | - | -0.024 | 0.002 | -0.011 | 0.016 |
| Severity |  |  |  |  |  |  |  |  |  |  |  |  |  |  |  |  |  |  |
| Mild | Ref. | - | Ref. | - | Ref. | - | Ref. | - | Ref. | - | Ref. | - | Ref. | - | Ref. | - | - | - |
| Severe | -0.004 | 0.970 | 0.422 | 0.196 | 0.189 | 0.220 | 0.149 | 0.195 | -0.032 | 0.904 | 0.087 | 0.434 | 0.023 | 0.849 | 0.379 | 0.144 | 0.109 | 0.451 |
| Critical | -0.013 | 0.935 | **0.804** | **0.092** | **0.364** | **0.073** | 0.198 | 0.243 | **1.014** | **0.004** | 0.207 | 0.203 | 0.048 | 0.789 | 0.494 | 0.191 | 0.246 | 0.195 |

Importantly, in the experiments reported here no SARS-CoV-2-specific antibodies were detected in more than 70 serum samples collected pre-pandemic. However, the majority of these pre-COVID-19 sera did contain antibodies against the nucleoprotein from the related HCoVOC43 betacoronavirus, that causes mild common cold-like diseases (Figure 6). Thus these data demonstrate that prior infection with another coronavirus does not seem to lead to the generation of antibodies cross-reactive with the SARS-CoV-2 virus.

**Figure 6.**
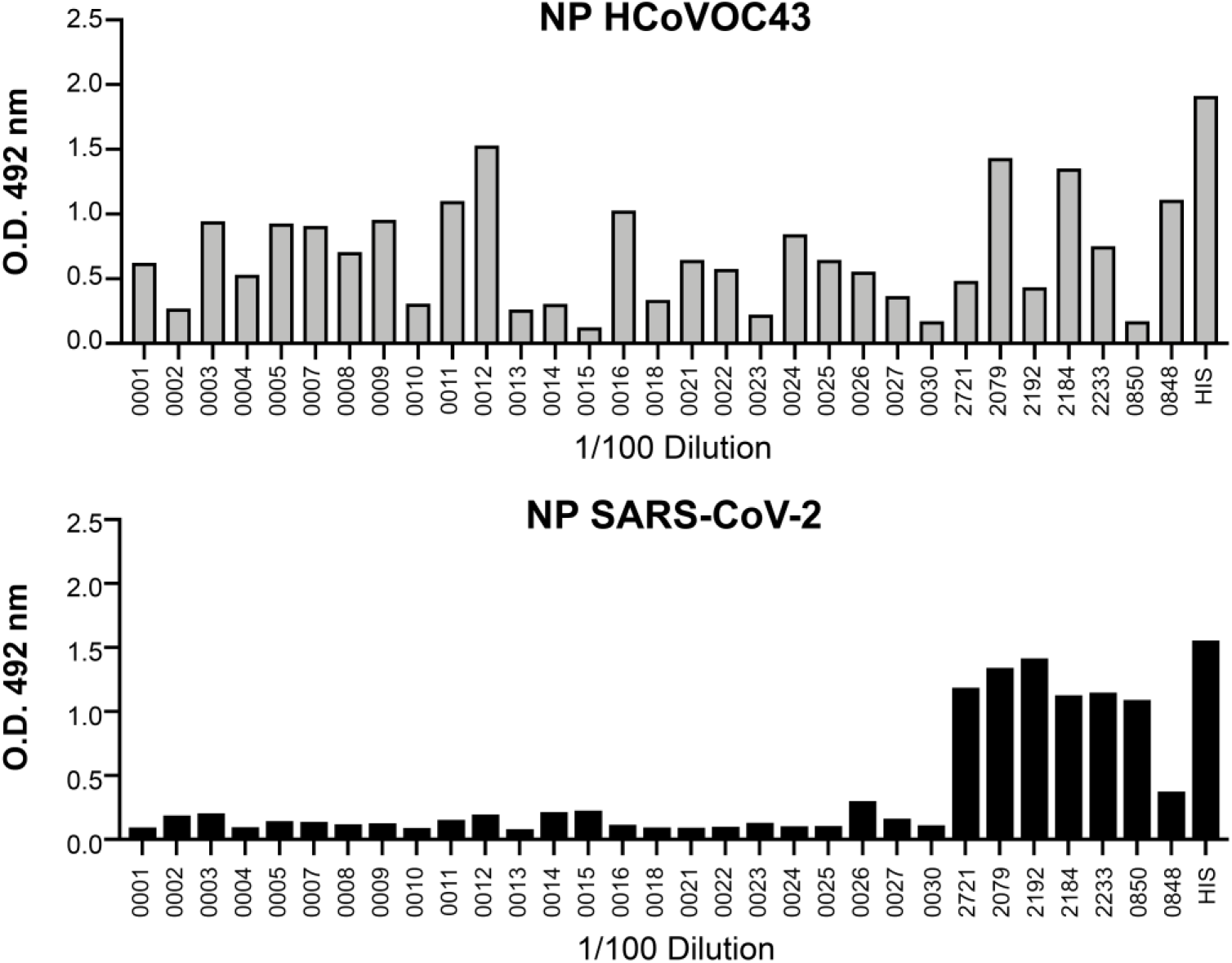
No cross-reactivity is observed between proteins from SARS-Cov-2 and OC43 betacoronaviruses. Plates were coated with 0.5 μg/ml of either SARS-CoV-2 NP or OC43 NP as indicated. Sera collected before 2020 (Pre-COVID-19) were tested at a 1/100 dilution. Detection was performed using antibody directed against human IgG. The bars labelled “2721-0848” correspond to COVID-19 PCR+ sera; the wells in which the amount of coated protein was tested by incubation with either anti-His are indicated. 13 out of 20 (65%) pre-COVID sera and 4 out of 7 (57%) COVID-19+ were clearly seropositive for OC43 NP. The donors with higher titres for OC43 anti-NP antibodies do not respond against SARS-NP, indicating that prior infection with OC43 does not lead to generation of antibodies reactive with SARS-CoV-2 antigens.

Therefore, the use of SARS-CoV-2 Mpro, in combination with other antigens already described for serology tests, provided outstanding specificity and sensitivity for patient identification. IgG titrated further than IgA or IgM indicating that, as expected, the IgG subclass is more abundant in serum. Assay for IgM antibodies had a lower signal/noise ratio and, in many of the SARS-CoV-2 negative sera a significant background could be observed for IgM. In contrast, SARS-CoV-2-specific IgA antibodies were not detected in healthy donors, but were clearly present in 27 out of the 36 sera tested from COVID-19 patients.

### Mpro-specific IgG antibodies are detected in saliva from COVID-19 patients

Saliva samples were collected from 11 healthy donors and 12 COVID-19 patients at the University Hospital La Princesa (Madrid) and tested in ELISA assays over a range of dilutions (1/2 to 1/10). IgG recognizing the three viral antigens tested could be observed in COVID-19 patients, with the strongest responses being those specific for the viral protease Mpro (Figure 7). IgA responses were detected in only one of the COVID-19 infected individuals (data not shown).

**Figure 7:**
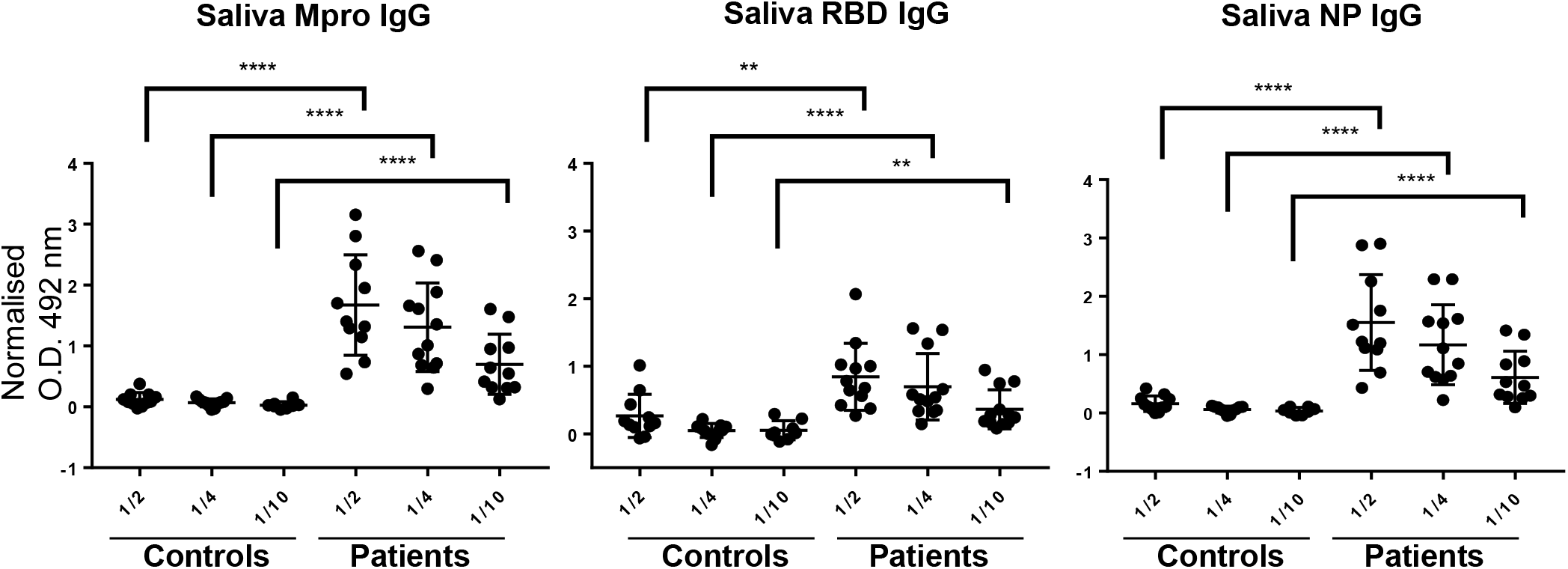
Comparison of saliva from healthy donors and 12 COVID-19 seropositive individuals. Plates coated with either 0.5 µg/ml of SARS-CoV-2 Mpro and NP or 1 µg/ml of RBD and ELISA tests were carried out on saliva samples diluted 1/2 to 1/10. Detection was done using antibodies directed against human IgG. Data were normalised for each antigen using the signal obtained for the positive control histidine-tag. Mann-Whitney test was performed to compare the values obtained for each dilution in healthy donors and patients. ** p<0.01, **** p<0.0001.

## DISCUSSION

The results presented here describe the detection of antibodies against the SARS-CoV-2 protease, Mpro, in serum from COVID-19 patients. The titres of Mpro-specific antibodies were comparable to those produced against SARS-CoV-2 nucleoprotein and somewhat higher than the antibody responses to the RBD fragment of the Spike glycoprotein, both of which are generally considered immunogenic coronavirus proteins. These high titre antibody responses in serum were accompanied by the detection of Mpro-specific IgG antibodies in saliva, providing a new opportunity for completely, non-invasive diagnostic tests.

For IgG antibodies in sera, the titres of NP and Mpro-specific antibodies correlate very well with each other (r=0.94 y p<10^−4^) and also with anti-RBD responses (r=0.89 y p<10^−4^). In contrast, while NP- and Mpro-specific antibody titres also correlate well for IgA and IgM responses (r values greater than 0.9), the correlation with IgM and IgA for RBD is much weaker (r values around 0.6). One plausible possibility is that the antibody responses to internal antigens, Mpro and NP, correlate well, since production of antibodies against these proteins requires either viruses with a broken membrane or release of viral material from infected cells.

The correlation with clinical data and symptoms onset reveals that antibodies have higher titres as the severity of the disease increases. Although the sample size is not large, this correlation was significant and independent of age and time from the beginning of symptoms for anti-RBD IgA and almost significant for anti-Mpro IgM and IgA. The retrospective design of our study does not allow to determine whether these increased levels are cause or consequence of more severe disease and what is the basis of its relationship with higher levels of IL-6 detected in critical patients. In this regard, it is surprising that IgM persisted at high levels in patients’ sera for more than a month after the beginning of symptoms.

The finding that the protease Mpro can be antigenic opens a new series of questions on the biology of this protein that is an important target for the development of antivirals to block SARS-CoV-2 replication. Mpro is key for cleavage and activation of the first polypeptide translated after infection, but the protein has not been found in the virion. So, most probably, the generation of antibodies directed against Mpro occurs at the end of the viral life cycle when intracellular antigens are released from the infected cell. It is not clear whether antibodies specific for Mpro might interfere with viral replication directly, however B cells producing these antibodies would likely efficiently internalise and present this antigen to stimulate T cell recognition of peptides from intracellular proteins.

The data presented here also show that, while antibodies for another betacoronavirus, HCoVOC43, were found frequently in pre-COVID19 sera, SARS-Cov-2-specific antibodies were undetectable, demonstrating that infection with one coronavirus does not necessarily prime for a better antibody response to another, at least for the viral antigens tested in these assays. Sequence analysis also suggests that it is unlikely that the response detected against NP and Mpro is due to cross-reactivity between coronavirus-specific antibodies. While COVID-19 Mpro has 96% homology with the main protease of SARS-CoV, which emerged in China in 2003, the similarity with other coronaviruses is much lower. All the samples analysed in this study came from hospitals in Spain, where no cases of SARS-CoV-1 have been reported. The similarity between the Cys-like proteases (Mpro) of different coronaviruses: SARS-CoV-2, HCovNL63, HCoVOC43 and HCov229E similarity is only around 40% with changes and similarities distributed along the whole sequence (Supplementary Figure 4).

A remarkable observation is that SARS-CoV-2 specific antibodies can be detected in the saliva of seropositive individuals. Two major antibody classes are found in saliva: secretory IgA (SIgA), synthesized locally by plasma cells (PCs) in salivary glands and IgG that is mainly derived from serum via gingival crevices (10). In our experiments salivary SARs-CoV-2 antibodies were mainly IgG rather than IgA; only one out of 12 individuals with SARS2-specific IgA was observed, corresponding to a donor that had recovered from the disease one month before the saliva test. The observation that COVID-19-positive, but not COVID-19-negative, individuals contain robustly detectable levels of SARS-CoV-2 NP and Mpro-specific antibodies in saliva is interesting because the development and validation of a saliva-based assay for SARS-CoV-2 seropositivity would represent a practical, non-invasive alternative to blood-based assays for COVID-19 diagnostic testing that might complement saliva-based nucleic acid tests for SARS-CoV-2 nucleic acid.

## METHODS

### Molecular cloning of the Cys-like protease (Mpro) and nucleocapsid (NP) proteins of SARS-CoV-2 and the NP of HCoV43

A gene encoding SARS-CoV-2 Mpro from the Wuhan-Hu-1 strain (ORF1ab polyprotein residues 3264-3569, GenBank code:MN908947.3) was amplified by PCR using the oligos 5’-gacccatggcttcagctgtttttcagagtggttt-3’ and 5’-gacctcgagttggaaagtaacacctgagcatt-3’, digested with NcoI and XhoI and ligated into the vector pET22b (Novagen) linearized with the same restriction enzymes.

Oligonucleotides 5’-gatccatggcttctgataatggtccgcaaaatcagcgtaatgca-3’ and 5’-caggtcgacaggctctgttggtgggaatg-3’were used to amplify the nucleocapsid protein of SARS-CoV-2. The amplification product was then digested with NcoI and SalI and ligated into the pET26b vector (Novagen) digested with NcoI and XhoI.

Oligonucleotides 5’-gatccatggtctcttttactcctggtaagcaatcc -3’ and 5’-gacctcgagtatttctgaggtgtcttcagtatag -3’were used to amplify the nucleocapsid protein of HCoVOC43. The amplification product was then digested with NcoI and XhoI and ligated into the pET26b vector (Novagen) digested with NcoI and XhoI.

The integrity of all constructs was verified by sequencing at MWG Eurofins.

### Expression of the SARS-CoV-2 Cys-like protease (Mpro) and nucleocapsid (NP) proteins

Recombinant viral proteins were expressed in the *E. coli* strain BL21 Star (DE3) pLysS (ThermoFisher).

SARS-CoV-2 Mpro protein was expressed by transforming this plasmid into the E. coli strain BL21 Star (DE3) pLysS. Transformed clones were pre-cultured overnight at room temperature in 50 mL 1 × LB medium with ampicillin (150 μg/mL) and chloramphenicol (34ug/ml). The overnight culture was then inoculated into 1L of 1 × LB medium (150 μg/mL ampicillin and 34ug/ml chloramphenicol) and the culture was grown at 37°C with agitation until the OD_600_ reached 0.6 when Isopropyl-D-thiogalactoside (IPTG) was added to 1mM to induce overexpression of the Mpro gene. The same protocol was followed to produce the nucleocapsid proteins except that kanamycin (150ug/ml) was used instead of ampicillin for antibiotic-mediated selection.

After overnight culture at 22°C for NP, 3h at 37°C for Mpro, bacteria were harvested by centrifugation at 9500 × g, 4°C for 15 min and the pellets were washed by resuspension in 150 mL TES buffer (20 mM Tris pH 8, 2mM EDTA, 150 mM NaCl) and re-centrifugation. Washed pellets were either processed immediately or stored frozen for later use.

Fresh, or thawed, cell pellets were resuspended in ice.cold 50 mM NaH_2_PO_4_ buffer pH8, 500 mM NaCl, 10 mM imidazole (I2399, Sigma Aldrich), 0.1% Sarkosyl, and 5% glycerol (pH 8.0). Lysozyme was then added (to 0.25 mg/ml) as were phenylmethylsulfonyl fluoride, Leupeptin and Pepstatin A (all to a final concentration of 1mM) and DNase I (2 µg/ml). Bacteria were lysed by sonication (3 cycles of 30 seconds with 30 seconds rest on ice between pulses) and soluble proteins were separated by centrifugation of the lysed cells at 14,000g at 4 °C for 45 minutes.

6-histidine tagged proteins were purified from the lysate using Nickel Affinity Cartridges 5ml (Agarose Bead Technologies S.L.). The bacterial supernatant was loaded on the column at a flow rate of 1 ml/min, followed by washing with 5 column volumes of 50 mM NaH_2_PO_4_ buffer, 500 mM NaCl, 10 mM imidazole and then 5 column volumes of 50 mM NaH_2_PO_4_ buffer, 500 mM NaCl, 25 mM imidazole. Recombinant proteins were eluted using a linear gradient of imidazole ranging from 25 mM to 250 mM over 5 column volumes (a representative SDS-PAGE analysis of the eluted fractions is shown in Supplementary Figure 1A). The proteins were then further purified by gel filtration using a 10/30 Superdex 75 Increase column (Cytiva) pre-equilibrated in 20mM HEPES, 1mM EDTA, 300mM NaCl, pH 7.5. The gel filtration analysis indicated that the SARS CoV 2 Mpro protein purified as a dimer.

### Molecular cloning and production of SARS-CoV-2 Receptor Binding Domain protein in mammalian cells (mRBD)

The cDNA region coding for the Receptor Binding Domain (RBD) (residues 334–528) defined in the structure of the S protein (PDB ID 6VSB) was amplified for expression in mammalian cells. The fragment was cloned in frame with the IgK leader sequence, an HA-tag (YPYDVPDYA) and a thrombin recognition site (LVPRGS) at its 5’ end, and it was followed by a second thrombin site, the TIM-1 mucin domain and the human IgG1 Fc region at the 3’ end. The recombinant cDNA was cloned in a vector derived from the pEF-BOS (11) for transient expression in HEK293 cells, and in the pBJ5-GS vector for stable protein production in CHO cells following the glutamine synthetase system (12). The inclusion of the TIM-1 mucin domain enhanced protein expression.

Mammalian RBD (mRBD) fused to the mucin domain and the Fc region (mRBD-mucin-Fc) was initially purified from cell supernatants by affinity chromatography using an IgSelect column (GE Healthcare). The mucin-Fc portion and the HA-tag were released from the mRBD protein by overnight treatment with thrombin at RT. The mixture was run through a protein A column to remove the mucin-Fc protein and mRBD was further purified by size-exclusion chromatography with a Superdex 75 column in HBS buffer (25 mM HEPES and 150 mM NaCl, pH 7.5). The concentration of purified mRBD was determined by absorbance at 280 nm.

### Baculovirus production of RBD-His tagged protein

A recombinant baculovirus expressing the RBD domain was generated using a pFastBac Dual-derived plasmid harboring the RBD coding sequence kindly provided by Dr. F. Krammer (6). HighFive (ThermoFisher Scientific) cell cultures were infected with the recombinant virus at a multiplicity of infection of 3 plaque forming units per cell and maintained in TC-100 medium (ThermoFisher Scientific) for 72 h. Thereafter, cell medium was harvested and clarified by centrifugation (4,300 × g for 10 min) and filtration through a 0.45 µm filter. Supernatant was loaded onto a chelated Nickel Affinity Cartridge-5ml (Agarose Bead Technologies S.L.) at a flow rate of 1.5 ml/min and eluted with a linear gradient of 500 mM Imidazole in Tris-saline buffer pH 7.5. Fractions were analyzed by SDS-PAGE and those containing RBD were pooled together and concentrated using an Amicon Ultra-15 Centrifugal unit with a 10 kDa cutoff membrane (Millipore). The concentrated protein was loaded onto a Superdex 75 10/300 Increase gel-filtration (GE Healthcare) equilibrated with PBS. The peak fractions were analyzed by SDS-PAGE and pooled together for further analysis.

### ELISA for detection of antibodies to SARS-CoV-2

96-well Maxisorp Nunc-Immuno plate were coated with 100 μL/well of recombinant proteins diluted in 0.1 M borate buffered saline (BBS) pH 8.8; NP and the protease at 0.5 μg/ml, RBD at 1μg/ml and incubated overnight at 4°C. Coating solutions were then aspirated, the ELISA plates were washed three times with 200 μl of PBS 0.05% Tween 20 (PBS-T) and then dried before blocking with PBS-casein (Biorad,1x PBS blocker) for 1 hour at room temperature. The plates were washed again with PBS-T and 100 μl of patient serum/plasma sample diluted in PBS-casein, 0.02% Tween-20, as indicated, was added and incubated for 2 hours at room temperature. The plates were washed again and 100 μL/well of the indicated detection antibody [(AffiniPure Rabbit Anti-Human IgM, Fcµ fragment specific; AffiniPure Rabbit Anti-Human Serum IgA, α chain specific; AffiniPure Rabbit Anti-Human IgG, Fcγ fragment specific) from Jackson Labs, or anti-human (Fab)’2 HRPO-labelled antibody from Thermo Fisher Scientific] was added and incubated for 1 hour at room temperature. The plates were washed with PBS-T four times and incubated at room temperature in the dark with 100 μL/well of Substrate Solution (OPD, Sigma prepared according to the manusfacturer’s instructions) (typically for 3 minutes). 50 μL of stop solution (3M H_2_SO_4_) were then added to each well and the optical density (at 492nm) of each well was determined using a microplate reader.

Negative controls included wells coated just with blocking buffer and serum samples collected from donors before 2019.

### Statistical analysis

Graphics and statistical analysis was performed with Graph Pad Prism 8 Software (GraphPad Software, USA, www.graphpad.com) and Stata 14.0 for Windows (Stata Corp LP, College Station, TX, USA). Quantitative variables following a non-normal distribution were represented as median and interquartile range (IQR) and the Mann Whitney test was used to test for statistically significant differences. Variables with a normal distribution were described by mean±standard deviation (SD) and differences between groups were assessed with Student’s t-test. Qualitative variables were described as counts and proportions and *χ*^2^ or Fisher’s exact test was used for comparisons. Correlation between quantitative variables was analysed using the Pearson correlation test.

Severity of COVID-19 was established as previously described (13). In this case, to determine differences in titres of antibodies between groups of severity the Cuzick’s test, that assesses trends across ordered groups, was employed.

Since several variables might contribute to differences in ELISA titres, multivariable linear analysis using generalized linear models (glm command of Stata) in which the dependent variable were ELISA titres of each isotype against each protein. The first model included age, gender and time from symptoms onset, followed by backward stepwise approach removing all variables with a p value >0.15 to obtain the best model for each protein and isotype. Then, the variable of interest (severity, anosmia or IL-6 serum levels) was forced in the model.

To determine the capacity of the different ELISA to discriminate between pre-COVID-19 sera and those sera obtained from patients with SARS-CoV-2, as determined by positive PCR from nasopharyngeal exudates, ROC analysis was performed, using the roctab command of Stata 14.1® (College Station, Texas). Each cut-off point was selected based on the best trade-off values between sensitivity, specificity and the percentage of patients correctly classified. ROC curves and area under curve (AUC) were also obtained.

### Patient samples and Institutional Review Boards

This study used samples from the research project “Immune response dynamics as predictor of COVID-19 disease evolution. Implications for therapeutic decision-making” [PREDINMUN-COVID] approved by La Princesa Health Research Institute (IIS-IP) Research Ethics Committee (register # 4070). Some experiments included patients from “Study of the lymphocytic response against SARS-COV-2, in different situations of host health and COVID-19 severity (InmunoCOVID)” approved by the Hospital La Paz Committee (HULP: PI-4101). All experiments were carried out following the ethical principles established in the Declaration of Helsinki. All included patients (or their representatives) were informed about the study and gave a written informed consent.

#### Patient selection

36 COVID-19 patients, diagnosed by PCR, were recruited for the study. 9 of them presented active infection by SARS-CoV2 at the moment of the study whereas the rest had no detectable levels of the virus. 10 patients required hospitalization, of which 6 were admitted to the ICU (Table 1). 33 serum samples from patients presenting a monoclonal gammopathy, allergic disease or rheumatoid arthritis, collected before June 2019 (PRE-COVID-19), were used as negative controls. All samples were stored frozen before use.

#### Antibody detection in saliva samples

12 donors with high antibody titres in serum were selected to measure specific IgG and IgA against SARS-CoV2 in saliva. For this purpose, new saliva samples were collected from these patients, and also from 11 healthy donors, aliquoted and immediately frozen. Prior to use, saliva samples were thawed, centrifuged at 400g and diluted 1/2, 1/4 and 1/10 in 1x PBS with 1% casein (Bio-Rad) and 0.02% Tween-20 supplemented with Complete™ Protease Inhibitor Cocktail (Roche).

## Data Availability

Data will be available on request. No large data set is included in this work

## Acknowledgements

The authors would like to thank the director of the CNB-CSIC, M. Mellado, for coordination; Luis Enjuanes and Sonia Zúñiga (CNB-CSIC) for SARS-CoV-2 DNA; Florian Krammer (Mount Sinai School of Medicine) for the plasmid for iRBD expression; César Santiago, Antonio J. Varas, Juan R. Rodríguez and José F. Rodríguez (CNB-CSIC) for the production and purification of iRBD. R. Delgado (Hospital 12 de Octubre, Madrid) for sera sample selection.

## Author contributions

HTR, DFS, GE, YCM, JMC, SP cloned and expressed proteins; JMRF, MVG, AA, PX, FSM, designed and optimized ELISA experiments; YCM, SG, AA, PM, TMA, LG, JMRF, MVG, implemented experiments; ELG, PM, AA, IG, FSM selected patients and performed clinical evaluation; IG, PM carried out statistical analysis; JMRF, MVG, FSM, HTR were responsible for the conception and design of the study and obtaining financial support; JMRF, MV, HTR, FSM, PM, AA wrote the manuscript with revisions from all authors

---

**Supplementary Figure 1.**
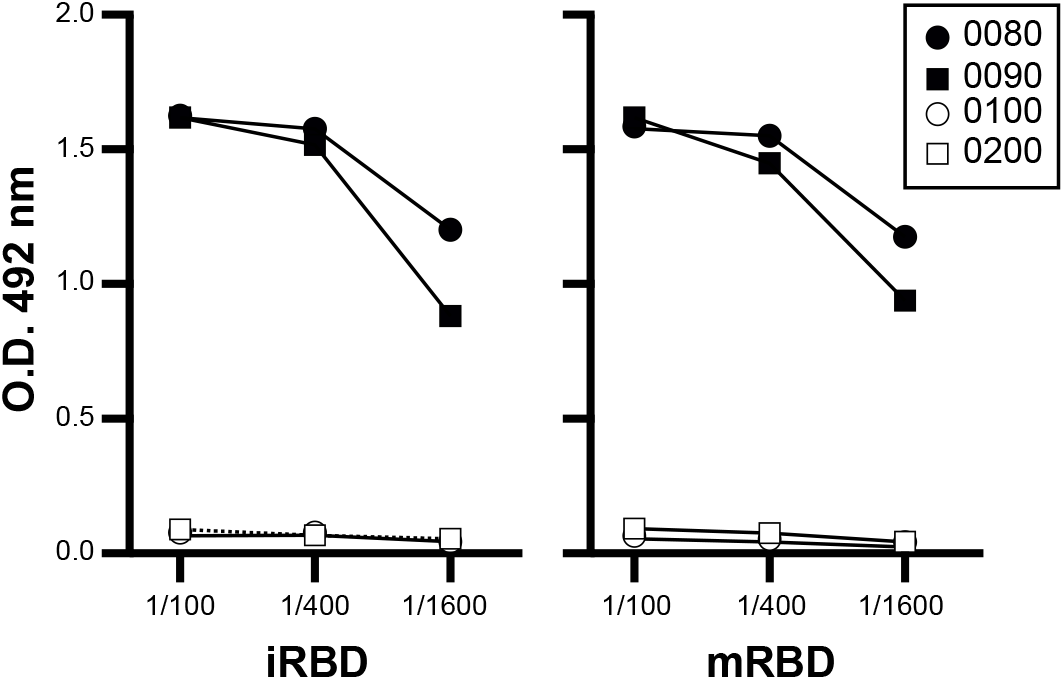
Detection of SARS-CoV-2 RBD-specific antibodies by ELISA. Plates were coated with SARS-CoV-2 RBD proteins produced in eukaryotic systems, either using insect or mammalian cells, and sera dilutions (1/100 to 1/1600) were tested. Detection was performed using anti-human IgG antibody. Black symbols correspond to COVID-19 patients and grey symbols to samples from donors pre-COVID-19.

**Supplementary Figure 2.**
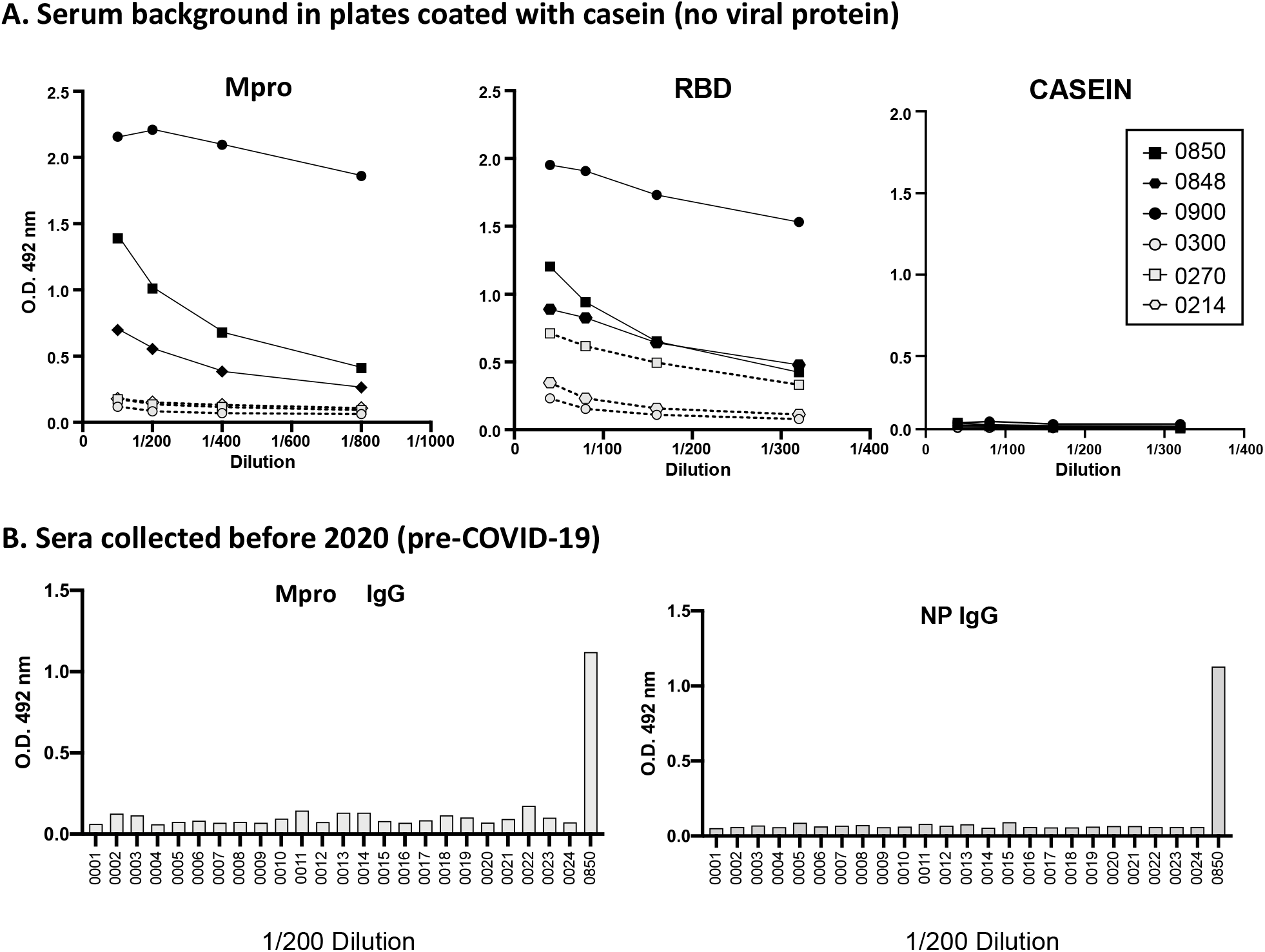
Background levels and negative ontrols. A. Background in plates with no viral protein coating. Plates were coated with 1 μg/ml of SARS-CoV-2 Mpro or iRBD and different dilutions of patient sera, as indicated, and detected with anti-human F(ab)2’ antibody (left and middle panels). Casein control corresponds to wells coated with the blocking solution, containing casein (right). These wells were incubated with the same sera and developed with anti-human F(ab)2’ antibody to check the background corresponding to individual sera. **B. SARS-CoV-2 negative controls**. 24 sera collected before 2020 (Pre-COVID-19) were tested in plates coated with 1 μg/ml of SARS-CoV-2 Mpro or NP. Sera were added at a 1/50-1/900 dilution. Detection was performed using antibodies directed against human IgG or IgM. Data from the 1/50 dilution are shown for IgM and 1/200 for IgG. Serum number 0850 corresponds to a positive control serum.

**Supplementary Figure 3:**
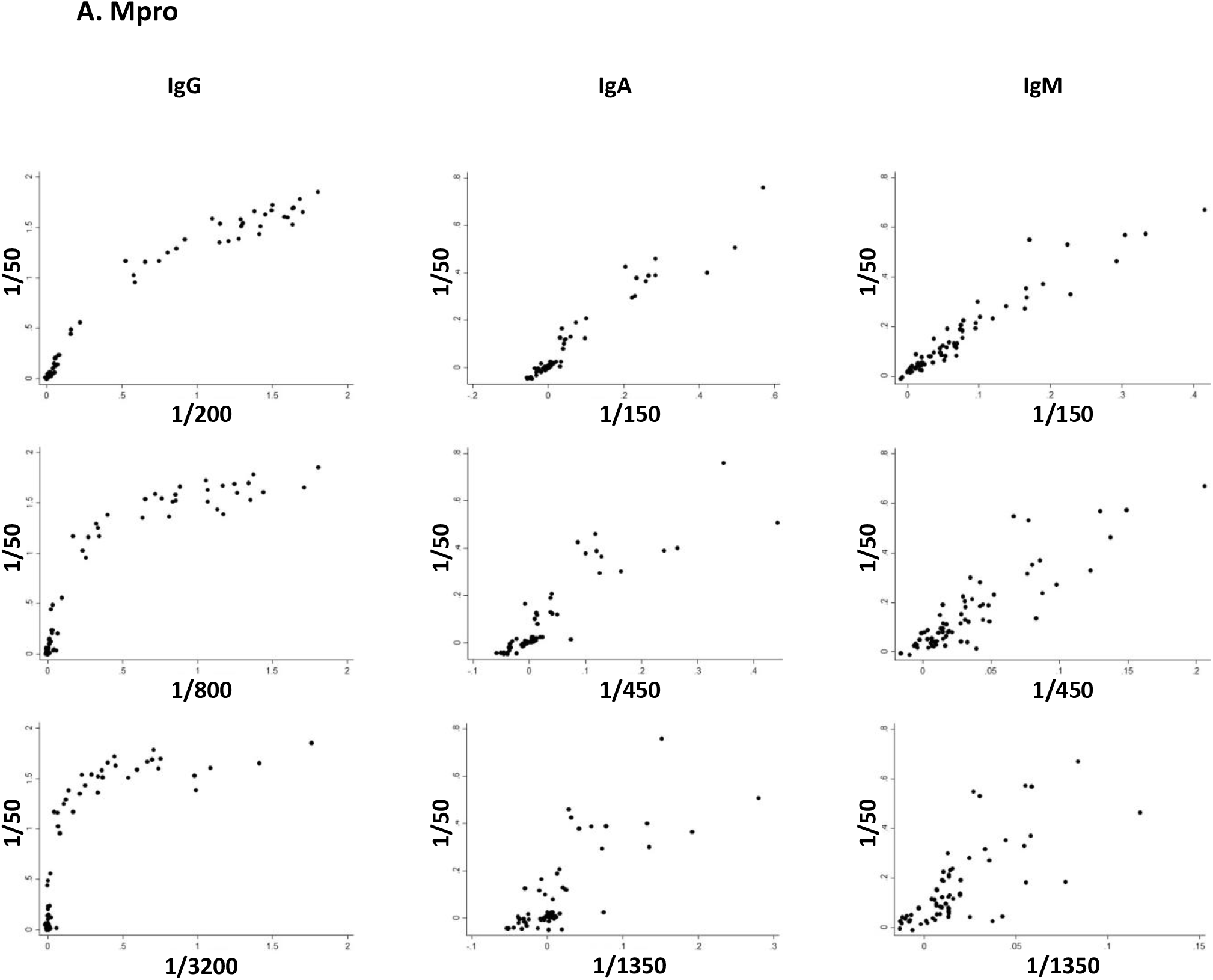
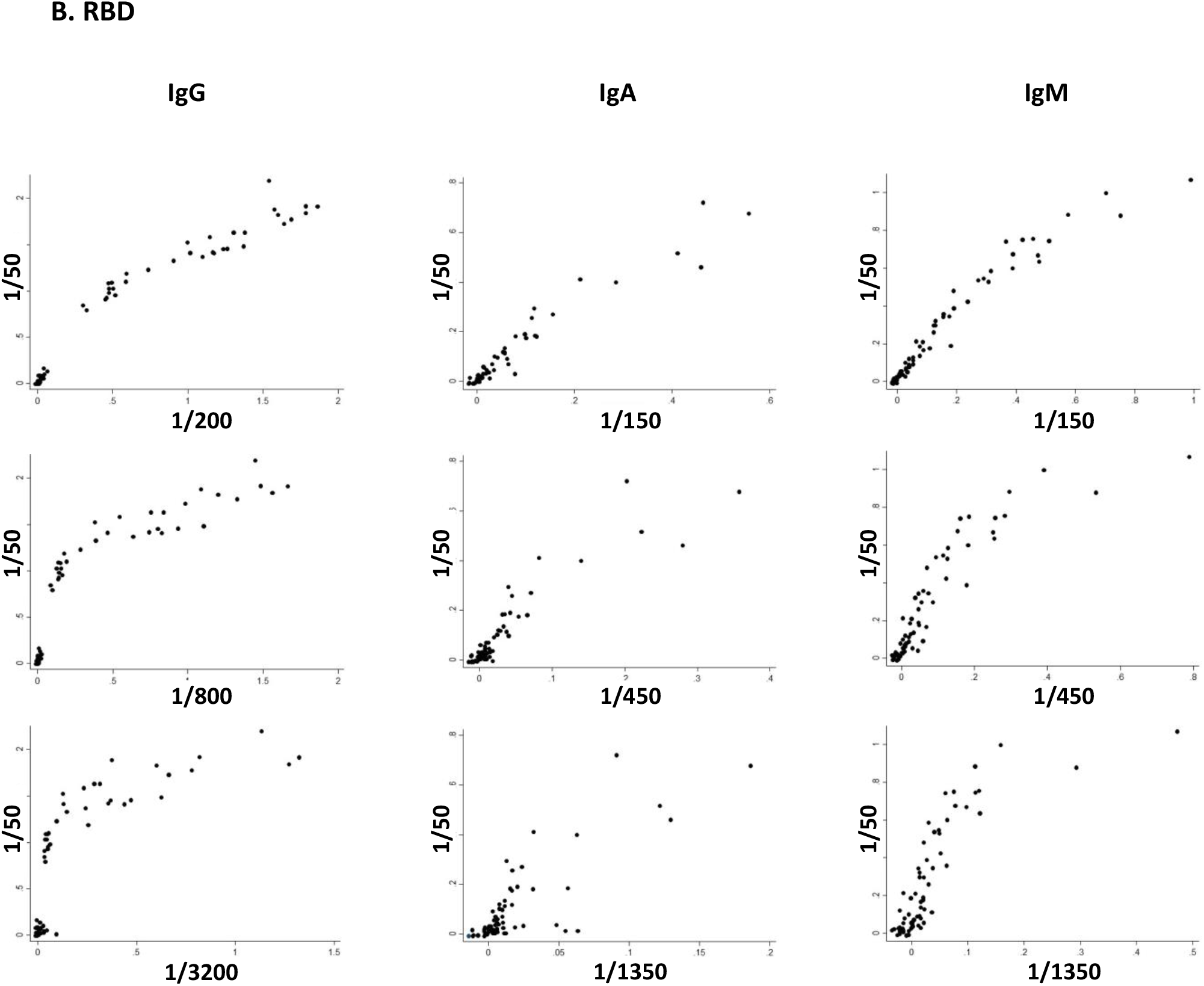
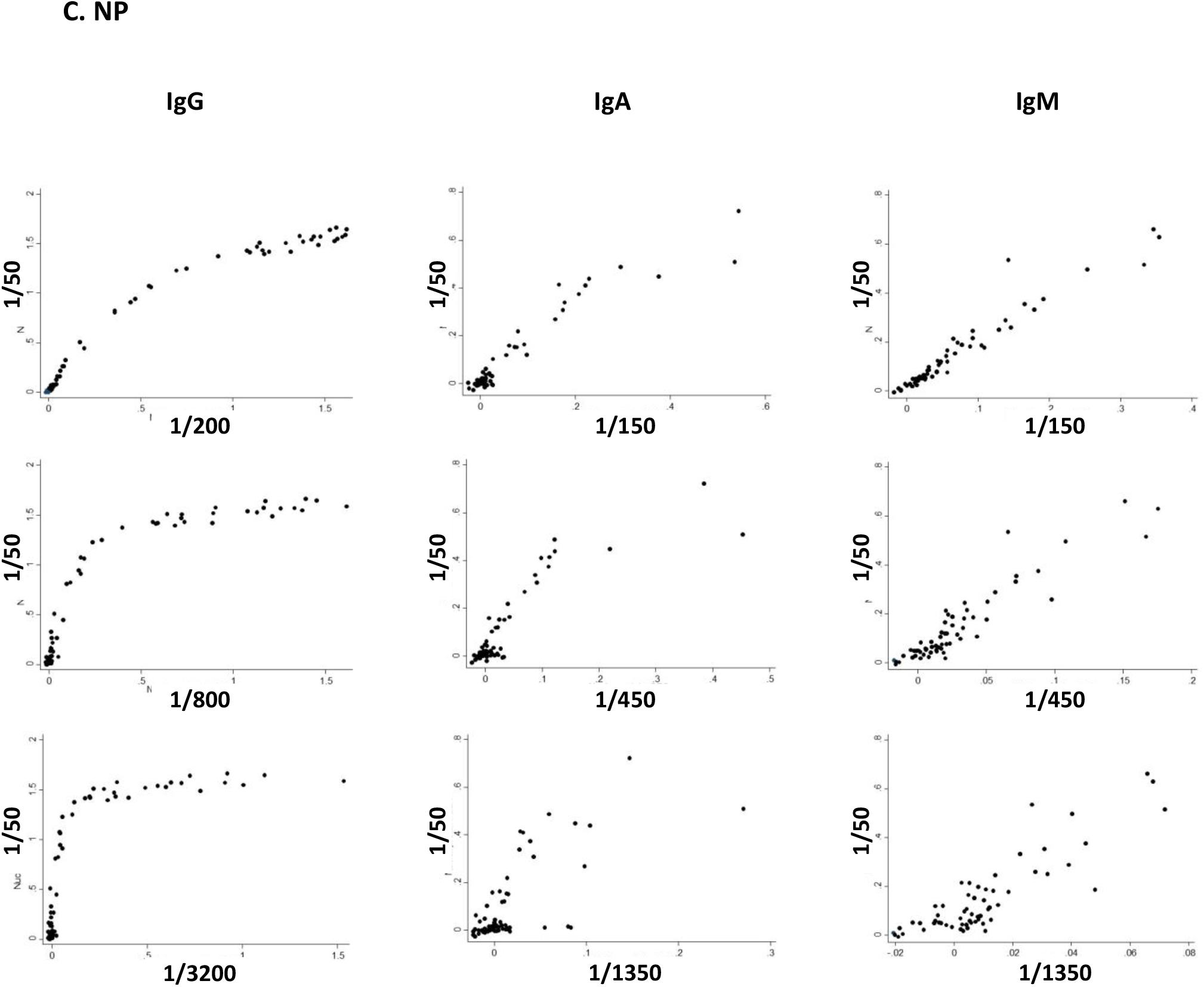
Comparisons between different sera dilutions for RBD, Mpro and NP. Plates coated with SARS-CoV-2 Mpro, NP or RBD were used to perform ELISA tests on 36 COVID-19 positive and 33 negative control sera. Detection was done using antibodies directed against human immunoglobulin of the three different subclasses: dilutions 1/50-1/3200 were used for IgG; dilutions 1/50-1/1350 were used for IgA and IgM. Graphs represent data of the ODs obtained for each antigen and each donor, after normalising the signal against a pool of positive sera. **A. Mpro. B. RBD. C. NP**.

**Supplementary Figure 4.**
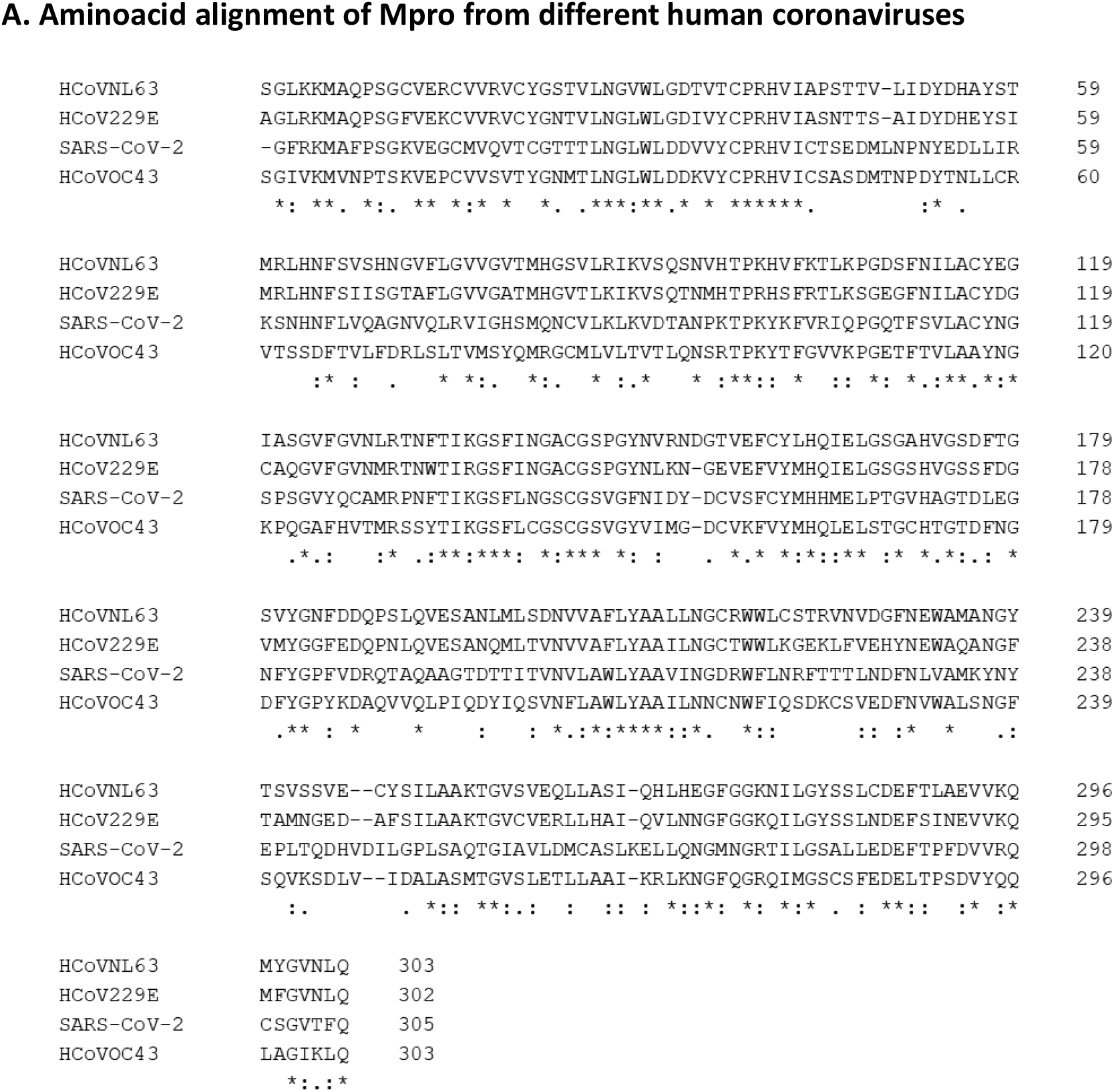
Alignment of Mpro amino acid sequences from the indicated coronaviruses. Sequences were obtained from the NCBI database and aligned using the Clustal Omega program (https://www.ebi.ac.uk/Tools/msa/clustalo/). * indicates positions which have a single, fully conserved residue, : indicates strong similarity,. indicates weak similarity.

## REFERENCES

1. Zhu N, Zhang D, Wang W, Li X, Yang B, Song J, et al. A Novel Coronavirus from Patients with Pneumonia in China, 2019. N Engl J Med. 2020;382(8):727–33.

2. Coronaviridae Study Group of the International Committee on Taxonomy of V. The species Severe acute respiratory syndrome-related coronavirus: classifying 2019-nCoV and naming it SARS-CoV-2. Nat Microbiol. 2020;5(4):536–44.

3. Liu R, Han H, Liu F, Lv Z, Wu K, Liu Y, et al. Positive rate of RT-PCR detection of SARS-CoV-2 infection in 4880 cases from one hospital in Wuhan, China, from Jan to Feb 2020. Clin Chim Acta. 2020;505:172–5.

4. Yu F, Yan L, Wang N, Yang S, Wang L, Tang Y, et al. Quantitative Detection and Viral Load Analysis of SARS-CoV-2 in Infected Patients. Clin Infect Dis. 2020.

5. Wang H, Li X, Li T, Zhang S, Wang L, Wu X, et al. The genetic sequence, origin, and diagnosis of SARS-CoV-2. Eur J Clin Microbiol Infect Dis. 2020.

6. Amanat F, Stadlbauer D, Strohmeier S, Nguyen THO, Chromikova V, McMahon M, et al. A serological assay to detect SARS-CoV-2 seroconversion in humans. Nat Med. 2020.

7. Bryant JE, Azman AS, Ferrari MJ, Arnold BF, Boni MF, Boum Y, et al. Serology for SARS-CoV-2: Apprehensions, opportunities, and the path forward. Sci Immunol. 2020;5(47).

8. Zhang L, Lin D, Sun X, Curth U, Drosten C, Sauerhering L, et al. Crystal structure of SARS-CoV-2 main protease provides a basis for design of improved alpha-ketoamide inhibitors. Science. 2020;368(6489):409–12.

9. Dai W, Zhang B, Jiang XM, Su H, Li J, Zhao Y, et al. Structure-based design of antiviral drug candidates targeting the SARS-CoV-2 main protease. Science. 2020;368(6497):1331–5.

10. Brandtzaeg P. Do salivary antibodies reliably reflect both mucosal and systemic immunity? Annals of the New York Academy of Sciences. 2007;1098:288–311.

11. Mizushima S, and Nagata S. pEF-BOS, a powerful mammalian expression vector. Nucleic Acids Research. 1990;18(17):5322-.

12. Casasnovas JM, and Springer TA. Kinetics and thermodynamics of virus binding to receptor. Studies with rhinovirus, intercellular adhesion molecule-1 (ICAM-1), and surface plasmon resonance. The Journal of biological chemistry. 1995;270(22):13216–24.

13. Wu Z, McGoogan JM. Characteristics of and Important Lessons From the Coronavirus Disease 2019 (COVID-19) Outbreak in China: Summary of a Report of 72?314 Cases From the Chinese Center for Disease Control and Prevention. JAMA 2020. doi: 10.1001/jama.2020.2648

